# Crohn’s disease and ulcerative colitis patient perspectives on clinical trials and participation

**DOI:** 10.1101/19000273

**Authors:** Orna Ehrlich, James Testaverde, Caren Heller, Stuart Daman, Annick Anderson, Peter D.R. Higgins

## Abstract

**Background:** Clinical trial recruitment is often the rate-limiting step in the development of new treatments reaching patients across all disease states. With more than 1500 currently available clinical trials for inflammatory bowel diseases (IBD) patients, it is important to understand patient perceptions of clinical trial participation to improve recruitment and retention. This study aimed to examine the specific challenges and barriers that might be reducing IBD patient enrollment and potential methods to overcome these barriers.

**Methods:** Five in-person patient focus groups were conducted from February through May 2016 using two facilitation guides. Participants self-reported a diagnosis of Crohn’s disease or ulcerative colitis.

**Results:** The five focus groups included a total of 34 participants. Participants discussed several barriers, including fears, disease severity at trial onset, potential adverse effects, time constraints, and the influence of both their primary IBD provider and support network. Methods to improve participation included better communication to prospective patients, reduced length of trial and time commitment, lower placebo rates, the option of open label extension, and support of the patient’s primary IBD provider.

**Conclusions:** This is the first study to examine patient perceptions for IBD clinical trial enrollment, including barriers to participation and methods to improve participation. Fear and misunderstanding of clinical trials, engagement with providers, limiting time demands, and limiting the impact on work and family were found to be barriers to participation. Creative solutions to these problems could lead to greater participation in trials and more rapid advancement of new therapies to clinical approval and use.

## Introduction

Clinical trial recruitment is often the rate-limiting step in the path to approval of new treatments for patients across all disease states. Slow recruitment causes delays in attaining the critical data needed to move the drug approval process forward. In fact, more than 70 percent of clinical trials are delayed at least one month because of slow enrollment^1^, which is costly for the trial sponsor as each day a drug is delayed from launch can cost sponsors up to $8 million in potential revenue^2^. In addition to the challenges of attaining a global recruitment target, more than half of the individual clinical research sites fail to meet enrollment goals in clinical studies^1^.

One cause of enrollment delay is failure to enlist the support of referring physicians to speak to their eligible patients regarding clinical trials. One study found that while the majority of people prefer to receive clinical research information from their primary care physician (51%) or their research team (44%), only a minority (23%) actually receive this information from them^3^. Reported reasons why providers do not inform their patients of available clinical trials are numerous and include a lack of awareness of actively enrolling relevant trials, reluctance to transfer care to another provider, lack of communication skills necessary to describe the protocol and clinical trial process to patients, and lack of time to present the information, among others^4^. Another cause of slow recruitment is the patients’ poor understanding of clinical trials, including the processes and safety protections built into modern clinical trials^1^. Ensuring that patients fully understand the importance of clinical trials and the role they play in making better therapies available to all is central to the success of increasing the number of therapeutic options for patients.

Inflammatory bowel diseases (IBD), which include Crohn’s disease and ulcerative colitis, affect 1.6 million Americans. On ClinicalTrials.gov there are close to 1,500 clinical trials currently available for Crohn’s disease and ulcerative colitis patients, but most of them will fail to enroll their target number of patients^5^. Understanding the specific challenges and barriers that reduce IBD patient enrollment, as well as finding ways to overcome these barriers, is imperative to improve recruitment and retention of patients in clinical trials. Thus, identifying and characterizing these barriers and potential solutions was the aim of this qualitative study organized by the Crohn’s & Colitis Foundation.

## Methods

Five in-person patient focus groups were conducted from February through May 2016. The Center for Information and Study on Clinical Research Participation (CISCRP), an independent non-profit organization dedicated to educating and informing all about clinical research and the role each party plays in the process, organized and facilitated the first two patient focus groups – one among patients diagnosed with Crohn’s disease and another among patients diagnosed with ulcerative colitis – in New York, NY in February, 2016. The results led to the revision of the facilitation guide based on initial feedback and the addition of three focus groups by the Survey Research Institute (SRI) at Cornell University, including Atlanta, Los Angeles, and Washington, DC in May, 2016 to provide geographic representation.

For all five sessions, participants in each group were recruited locally by the Crohn’s & Colitis Foundation and were composed exclusively of IBD patients (by self-report). The Foundation recruited participants through various channels, including email invitations to constituents in the local area and through social media. All potential participants were then asked demographic questions and provided more information on the focus group by phone with a Foundation staff member. IRB exemption was received for all focus groups and patients were consented at the beginning of the session. The sessions were audio recorded and transcribed. Focus groups were conducted in a conference room in a non-medical building to avoid bias in location. Questions were largely open-ended and posed in a non-leading fashion; if no one responded, then the facilitator used prompts to stimulate discussion. Additionally, the facilitator made it clear at the start of the discussion that they were from an independent organization to encourage open feedback.

The focus group facilitation guides were composed of a series of questions about clinical trials. Questions fell into several categories: impressions and knowledge of clinical trials, learning about clinical trials, past experiences (if any), education to decide whether to participate in a trial, factors that would influence the decision, and expectations of participation. The base questions were determined through CISCRP experience with past patient advisory boards, as well as learnings from the global CISCRP perceptions & insights study. A full facilitation guide is presented as an appendix to this manuscript.

The transcripts from each focus group were saved as a plain text file and edited to remove the names of speakers, organizing and closing statements, and any statements by the interviewers. The remaining 1946 blocks of transcribed text were then isolated as 88,686 words with the use of the R package *tidytext* by Julia Silge and David Robinson. These words were analyzed for sentiment, using the NRC Word-Emotion Association Lexicon of Saif Mohammad and Peter Turney and the R package *syuzhet* by Matthew Jockers. Additionally, a word cloud of commonly used terms was created. A network plot of two-word bigrams was created with the *ggraph* R package by Thomas Lin Pedersen, available from the CRAN repositiory (https://cran.r-project.org/web/packages/ggraph/index.html).

CISCRP and SRI also used a thematic analysis approach to identify and code themes through a review of the transcripts. The themes included those topics, concerns and sentiments that were interpreted as more important to patients. This included being discussed by patients at multiple focus group sites, by more patients within individual sites, or in response to multiple questions throughout the discussion. The breadth and diversity of sentiments were also captured in the themes. Themes were hand-coded across transcripts (no software used).

## Results

The five focus groups included a total of 34 participants (21 females, 13 males). Diagnoses were evenly split between Crohn’s disease and ulcerative colitis (Table 1).

**Table 1.**
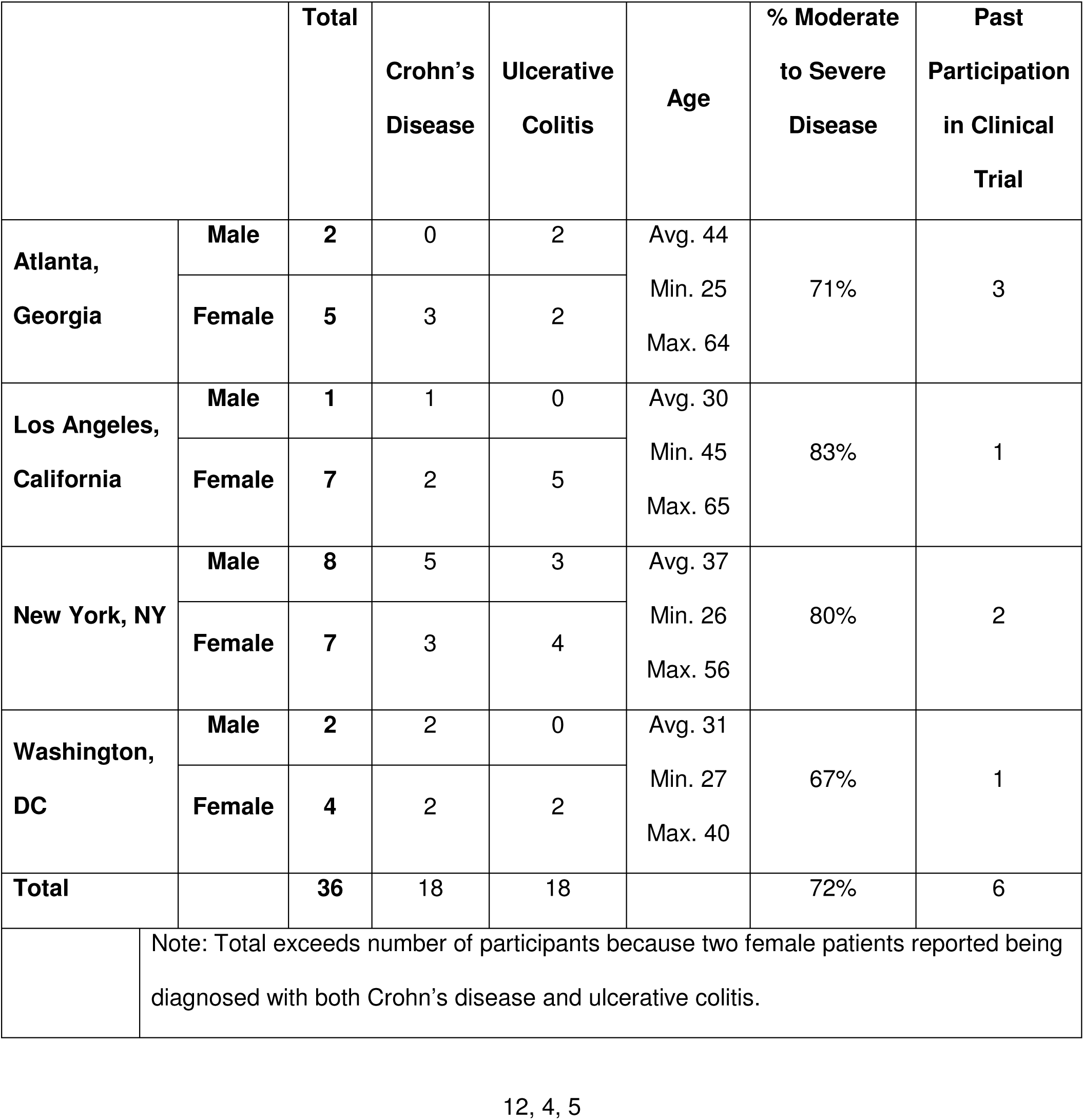
Distribution of IBD diagnoses in patients

Six patients (2 males) reported having been diagnosed with IBD less than five years ago, ten patients (6 males) between five to ten years ago, seven patients (2 males) between 10 and 20 years ago, and 10 patients (2 males) were diagnosed more than 20 years ago. Among those able to recall their age of disease onset, the youngest was 12 and the oldest was 44.

### Impressions and knowledge of clinical trials

All patients in the sample claimed to know what clinical trials are at a basic level. However, many misconceptions and questions remained, reinforcing the need for IBD patient education on clinical research. For instance, several patients mentioned being more likely to consider a clinical trial when they are less sick, which indicates a lack of understanding of the parameters for clinical trial eligibility as individuals on effective treatments would not be enrolled. Most patients claimed that they knew or were pretty sure they knew the distinction between Phase 2 and Phase 3 trials. Some patients in the latter focus groups felt they had some understanding of the difference between induction, maintenance, and open label extensions, but that they were not necessarily familiar with the formal definition of these terms or concepts.

Patients reported being attracted to clinical trials because of the potential to help people, including themselves, in the present or future. Some also emphasized the hopeful nature of having more current and future options for medications, as they were aware that any one medication does not work for all IBD patients. Common concerns about IBD clinical trials were the fear of being assigned to a placebo group, the unknown efficacy and potential side effects of the drug, the negative impacts that trial participation could have on other aspects of their lives (e.g., child bearing), and confusing advertising for trials.

Most patients said they would seek information about trials from their doctor, patient support groups and by searching the internet. Several patients suggested that they would benefit from hearing from other patients who had participated in a clinical trial. Doctors, patient support groups, and websites were suggested as trusted source, while traditional media (e.g., TV and radio commercials) was generally described as an untrustworthy source.

When focus group subjects discussed clinical trials, their dominant sentiment was anticipation of new and better treatments, with a relatively high degree of trust. The sentiment analysis scores in 10 dimensions are presented in Figure 1. Negative sentiments, including disgust, anger, fear, and sadness, were common. Among positive sentiments, surprise was common, and joy was uncommon in these focus groups. A word cloud displaying the most commonly used words in the five focus groups is presented in Figure 2. The centrality of doctor, time, work, and information reflect several of the most common concerns of patients about clinical trials in IBD. A network plot of the words used in pairs in the focus groups is presented in Figure 3, and illustrates the perception of the intersecting interests of insurance companies and pharmaceutical companies, and the clustering of time and travel concerns.

**Figure 1.**
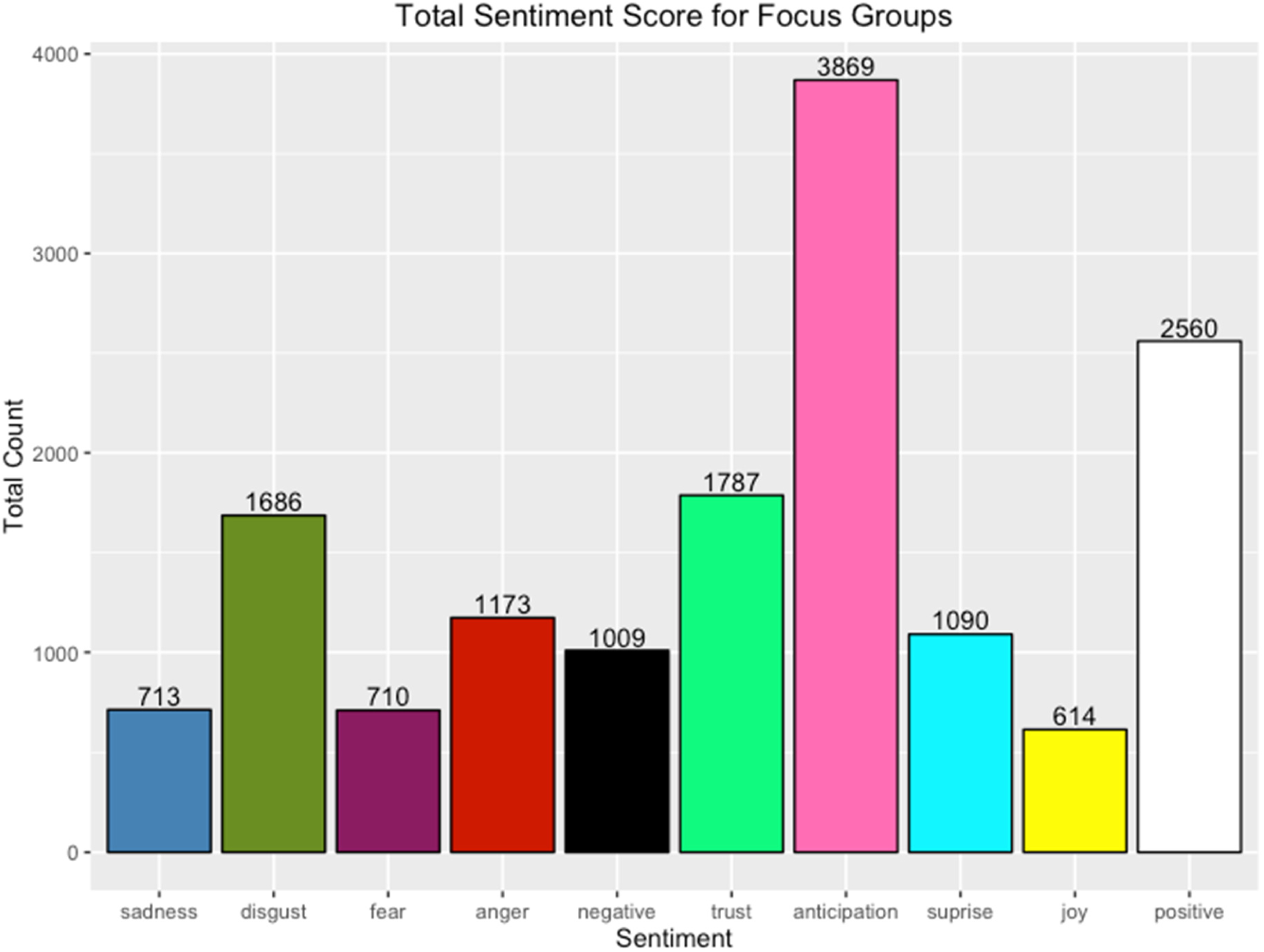
IBD Focus Group sentiment analysis, with words coded to sentiment using a standard English language lexicon. Height of bars indicates the count of words with each sentiment.

**Figure 2.**
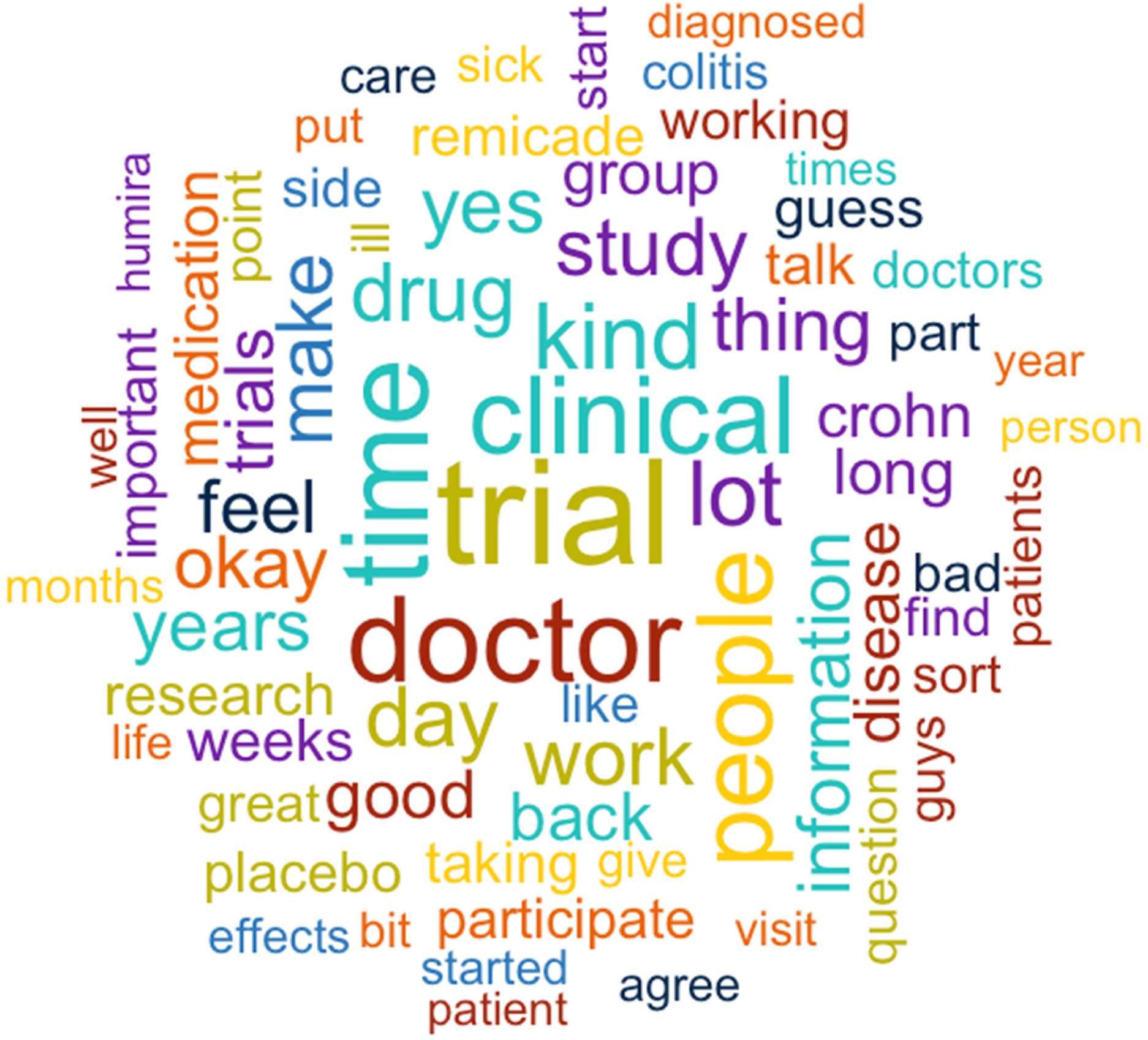
IBD Focus Group word cloud, with more frequent words represented as larger and closer to the center. Colors are only for contrast.

**Figure 3.**
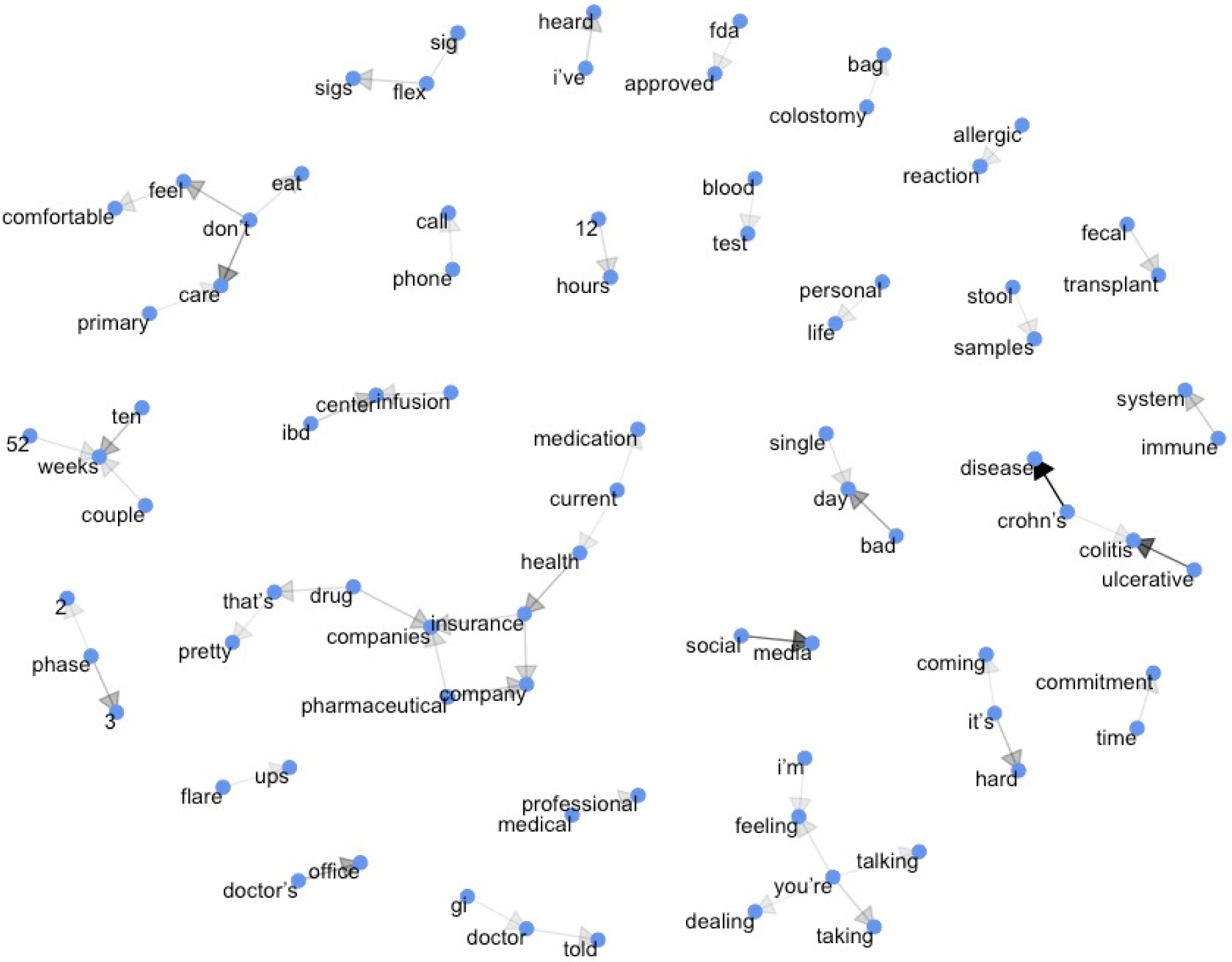
IBD Focus Group network plot of words used together. Interesting interactions include the intersection of insurance companies and pharmaceutical companies in the lower left, time commitment and travel to sites in the lower right, and stool samples and fecal transplant at the upper right. Arrows indicate the sequencing of the words, and the darkness of the arrows the frequency of the sequence.

### Experiences participating in clinical trials

Of the total 34 patients, nine (26%) reported having been involved in an IBD clinical trial previously. Former trial participants were present in each of the five live sessions. Patients’ motivation for participating mostly revolved around the possibility of improving their health (therapeutic misconception). One patient described being in a particularly severe IBD flare, and being more willing to try a new drug that might work. Another was told that remission could come faster with a new therapy, and was motivated by this. Two other patients were specifically motivated by the price of the (free) medication involved.

The most frequent negative comments regarded dislike for processes or procedures that were part of the trial, including long wait times for laboratory visits or pharmacies, lengthy surveys, and other tasks required to document trial endpoints (e.g., keeping a diary or making a video). Other negative comments included discomfort with getting multiple colonoscopies in a single year and fear of negative unexpected side effects that researchers were not and could not be aware of. The six patients from the three latter focus groups who had prior trial experience were asked whether they would be willing to participate in another clinical trial if they qualified, and all said yes and generally had positive comments regarding their overall participation.

### Learning enough to decide to participate in a trial

Some of the discussion focused on how patients would gather information about a new trial in order to make a decision about participation. In order to make a decision about whether or not to participate, patients said they would want a lot of information about the trial, including any available information on the study drug itself, including: results from earlier phase studies, expected time to study drug response, data on efficacy and side effects, use in other indications or therapeutic areas, number of patients already enrolled, and the total number that would be enrolled. They also wanted to learn about the types of procedures to expect as part of the trial and frequency of these procedures. Some wished to know more about the background of the drug, such as its mechanism of action. Others wanted to know who the sponsor for the trial was, and the specific goal of the trial. Patients were also interested to learn which medications were permissible in addition to the study drug during the trial as well as any open-label extension options (if available). Comprehensive care was also mentioned as important in the initial two focus groups, with options such as having access to a nutritionist, patient support groups, and the opportunity to speak with prior study volunteers to learn about their experiences.

#### Influence of provider and support network

A recommendation to participate in a clinical trial from a doctor they trust would be a strong motivator to participate in a clinical trial, and perhaps the most powerful single factor. Regardless of other factors, patients said that having the support of their primary gastroenterologist (GI) was important, with an average rating of 8.5 on a 1-10 scale (10 = most important) as summarized in Table 2. Participants valued their long-term relationships with their doctors and felt that their doctor would be in the best position to assess whether a particular clinical trial would be a good option for them, since their doctors were familiar with the course of their IBD.

**Table 2.**
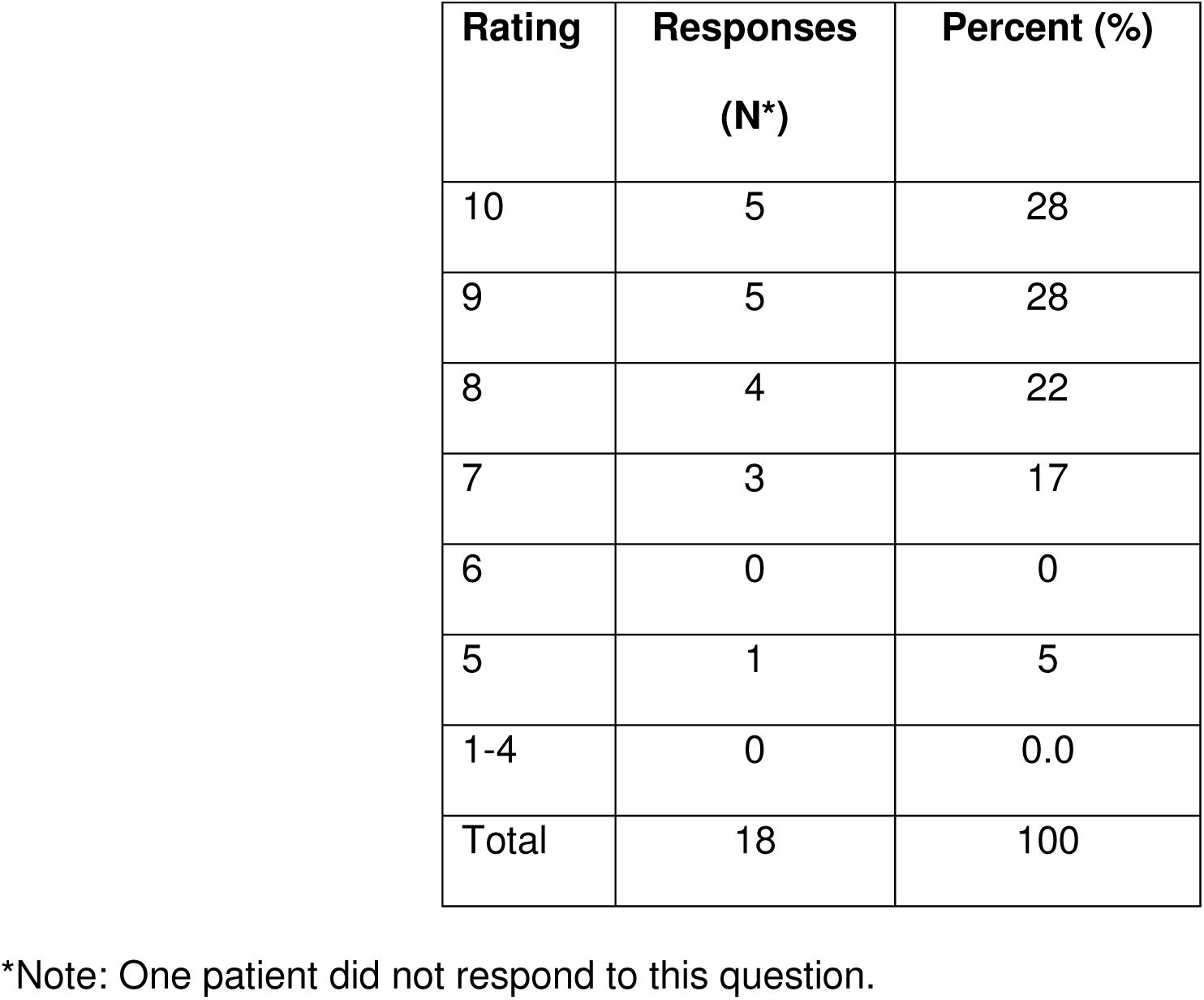
Frequency distribution of ratings of importance of support of primary gastroenterologist among three focus groups

Additionally, the support of family was important, and concerns of family members could be a barrier to participating. Patients reported wanting support for making the specific decision to participate in a trial. Most often, this support should come from family, whether it was parents, spouses, or children. Patients also preferred to have approval from their doctor and very few patients said that *disapproval* of their doctor would *not* be a barrier to participating in a trial.

### Important factors to determining attractiveness of trials

#### General Factors

When asked what determines whether trials are attractive to patients, the focus group members described many criteria. First, patients would not feel attracted to trials if their current health was good, stable, or under control (i.e., with their current medication regimen). Second, patients expressed much less interest in trials that required a large time commitment, whether that meant long individual appointments or duration of the entire trial, or the time required to travel to the trial location. Other important factors included the lifestyle changes that would result from participating in trials, such as the impact on time with one’s family and time taken away from work.

#### Placebo

Patients had strong feelings about the presence of placebos in clinical trials. A few patients conveyed the understanding that getting a placebo was an inherent risk of being in trials and explained that it might not be a good experience, particularly if their condition worsened during the trial, but they would still be willing to enroll with this risk in mind. On the other hand, several patients said that knowing they would get a placebo or have a higher chance of receiving one would make them less interested or not willing to participate. Several wanted reassurance that a rescue option would be available if they were doing poorly during the induction phase, and that a flare and rescue while on placebo would not disqualify them from eventually receiving the investigational therapy. In general, a 20% chance of receiving a placebo seemed tolerable. A few patients elaborated on balancing this percentage with other factors, such as their current health or the time and travel burden associated with participation. An increase in other burdens would make them less likely to participate in a trial with a higher placebo rate.

#### Open label extension

Focus group participants found the option of potentially continuing on the study medication after the study ended attractive, but were concerned about the length of a trial before the open label option became available. The participants reported a range of periods of time they would be willing to continue on the study medication while feeling poorly before dropping out of trials. Most patients said they’d be willing to wait a while before dropping out, such as one, three, or six months, or half of the duration of the trial. The dominant thought was that they would drop out sooner if their health was not good or if it rapidly worsened. Patients emphasized that changes could happen fast, such that the decision to stop a trial medication could also come quickly, and some form of rescue therapy should be available for this situation. Thus, communication on the length of time a trial medication could take to induce a response would be important to set expectations for participants. A couple of participants also said that their current life circumstances would also matter, such as child care or elder care responsibilities, or their ability to get time off work. If a sponsor were able to help with these sorts of difficulties, patients said they would be more able and willing to enroll and remain in clinical trials.

#### Influence of location of trials

All patients said that the location of trials would be important. For comparison, a few patients conveyed that they were already traveling a moderate distance to reach their current doctor, because they believed their doctor was worth the travel time. Similarly, confidence in a medical facility where they would be participating in trials was considered worthy of greater travel. For example, facilities that conducted more trials, such as larger academic medical centers, were more appealing to patients. Another patient said that the doctor conducting the trial mattered more than the medical center, such as his or her familiarity and experience conducting clinical trials. One patient also mentioned that accessing such facilities could be more difficult for patients in rural areas.

When discussing the actual distance they would be willing to travel, most patients emphasized that the time they would travel was the greatest factor, whether they would have to travel during rush hour, and how frequently they would have to miss work to participate in the clinical trial. Specific time varied by location. For example, patients from DC said that two hours was the maximum they would travel, whereas patients from Atlanta said that 30-60 minutes was “not problematic”.

#### Influence of trial duration and frequency of visits

The duration of trials matters to patients, where shorter trials are more appealing (Table 3). Reasons for willingness to participate in trials of longer duration focused on three issues. First were concerns about having to take a lot of time off work for appointments and/or greater burdens on one’s family. The second issue was the concern that some medications might take a long time to achieve effectiveness, and if these expectations were made clear at trial entry, patients would be more willing to participate in longer trials of these types of drugs. The third issue they brought up was the concern about being in a placebo group; if there were a placebo arm, the participants reported that they would not want a long induction phase (“initial” period), and would want the opportunity for early exit and rescue therapy with the investigational therapy if they flared. The potential duration of a placebo arm was another factor that weighed into willingness to participate in a trial: longer studies with relatively high percentages in the placebo arm would be less appealing to patients, though this can be mitigated by the opportunity to exit to rescue therapy if a severe flare occurs. Participants also were concerned about the total time at each study visit. In the first two focus groups, the consensus was that generally 2 hours should be the maximum visit time with a few exceptions for longer visits. Additionally, scheduling flexibility was also found to be important in these first two focus groups, such as being able to make appointments after work hours or on the weekend, and being able to reschedule appointments when needed.

**Table 3.**
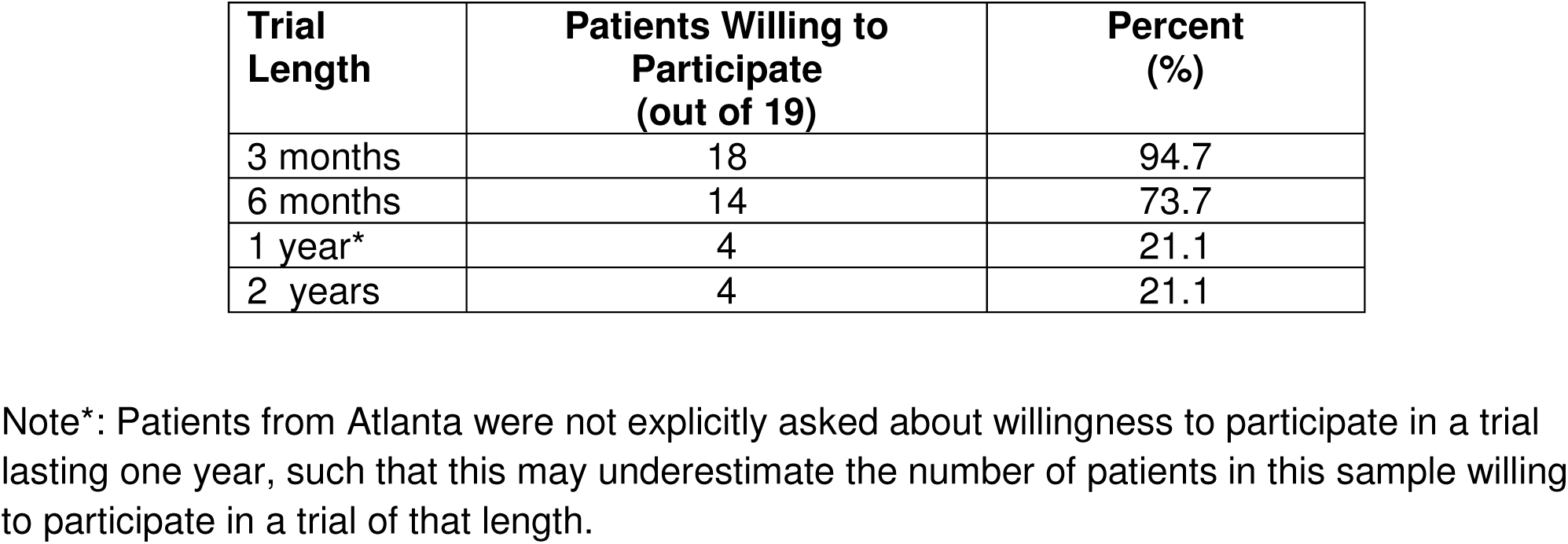
Frequency distribution of willingness to participate in trials of different durations among three focus groups

### Expectations of participation

Patients expected a high level of professionalism, customer service, and compassion at the facility where they would participate in a trial. A few expressed an expectation for a higher quality or level of treatment, because they were doing the sponsor a favor by participating.

Participants also described many expectations for what would happen after trial completion. Many expected follow-up, whether it would be multiple appointments over the following weeks or months, or just one appointment, before transitioning treatment back to their regular doctor. Several expressed a desire to know what the results of the trial were, such as whether the drug was determined to be effective or not. Two patients expressed a desire for post-trial support, such as a support group or another way to connect with other participants to compare side effects and other experiences.

## Discussion

Several themes emerged from the content review of the five focus group sessions on barriers to participation in IBD clinical trials and suggested several opportunities to improve clinical trial participation (Table 4). There were significant knowledge gaps about clinical trials for these participants, despite their claims to the contrary. This was not surprising, as a National Cancer Institute study found that 40% of adults lacked a basic understanding of how clinical trials were conducted^4^. Another study found that while self-reported general knowledge of clinical research was high (81% of adults claimed to be informed on clinical research with no significant differences across therapeutic areas)^3^; a different study discovered that most adults (63%) were unable to name a living scientist nor were able to accurately name a place where clinical research was conducted (64% did not know) ^6^. These studies provide further evidence of surface level knowledge on clinical research among a broader audience and highlight the importance of educational efforts.

**Table 4.**
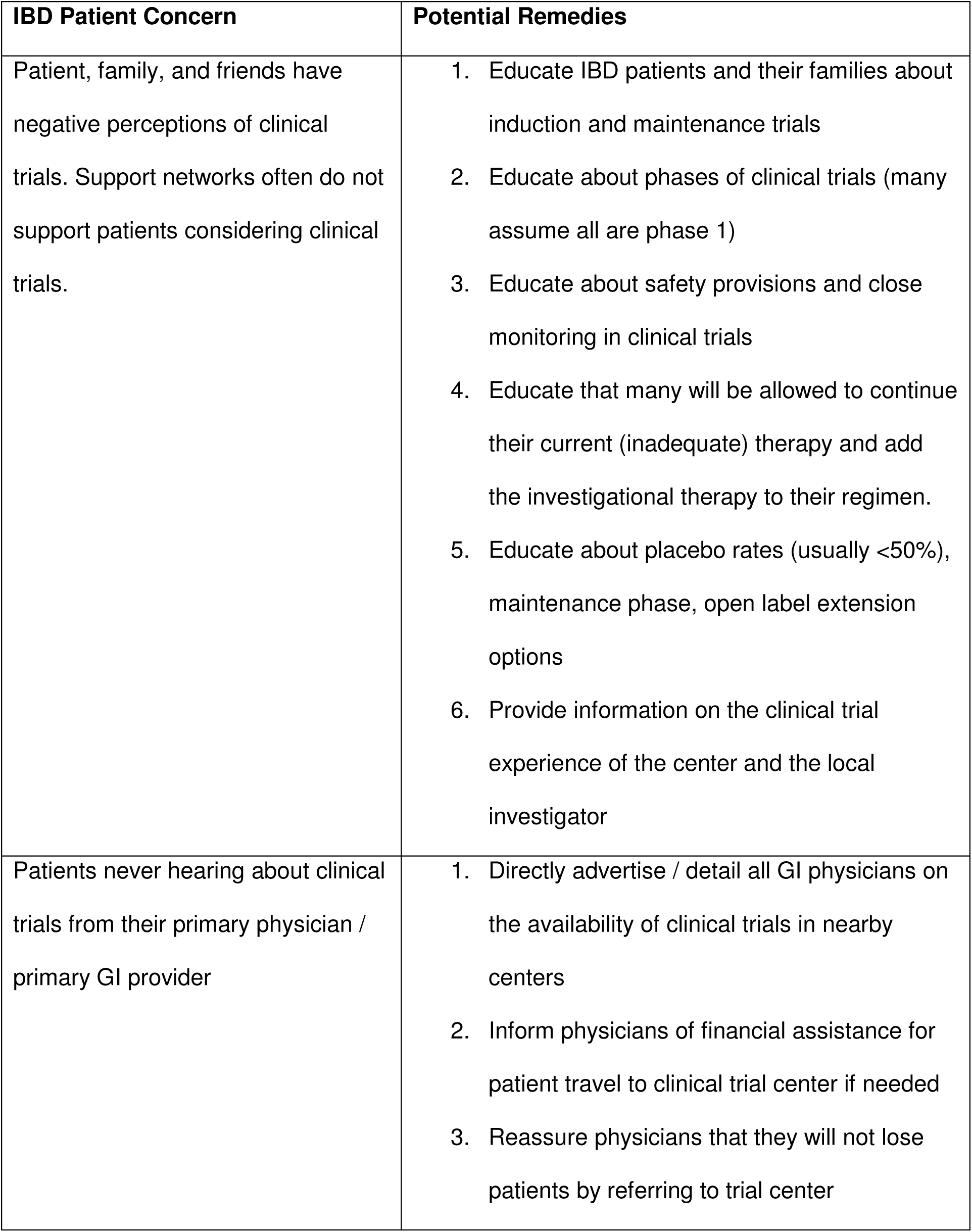
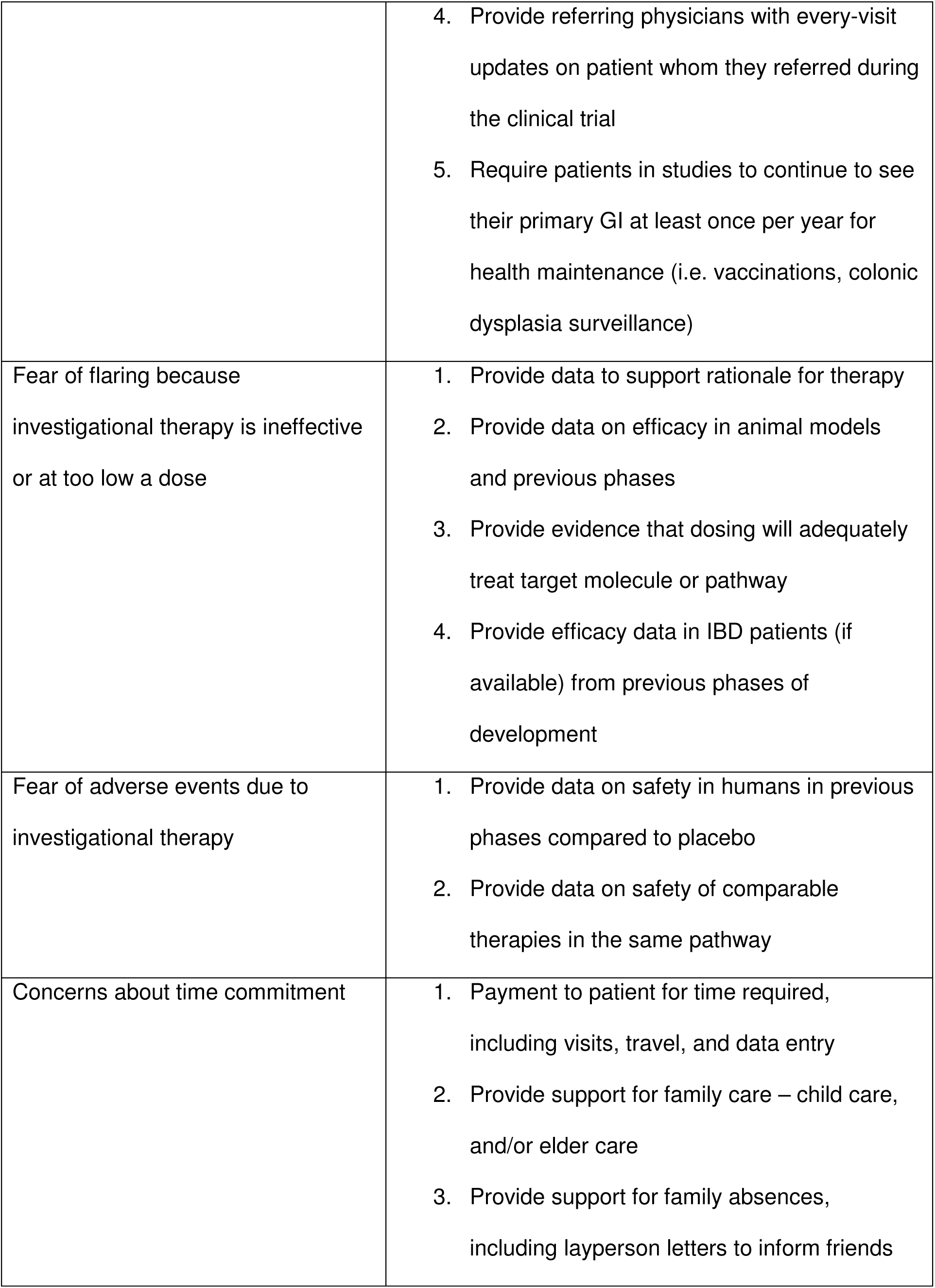
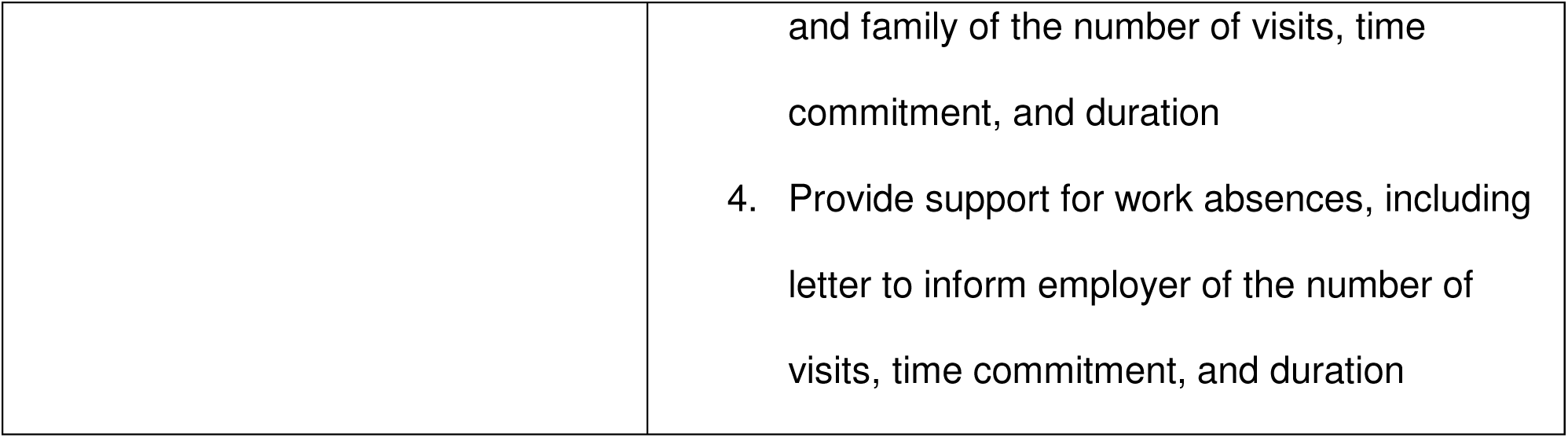
IBD patient concerns about clinical trials and potential remedies

| IBD Patient Concern | Potential Remedies |
| --- | --- |
| Patient, family, and friends have negative perceptions of clinical trials. Support networks often do not support patients considering clinical trials. | 1. Educate IBD patients and their families about induction and maintenance trials<br>2. Educate about phases of clinical trials (many assume all are phase 1)<br>3. Educate about safety provisions and close monitoring in clinical trials<br>4. Educate that many will be allowed to continue their current (inadequate) therapy and add the investigational therapy to their regimen.<br>5. Educate about placebo rates (usually <50%), maintenance phase, open label extension options<br>6. Provide information on the clinical trial experience of the center and the local investigator |
| Patients never hearing about clinical trials from their primary physician / primary GI provider | 1. Directly advertise / detail all GI physicians on the availability of clinical trials in nearby centers<br>2. Inform physicians of financial assistance for patient travel to clinical trial center if needed<br>3. Reassure physicians that they will not lose patients by referring to trial center |
|  | 4. Provide referring physicians with every-visit updates on patient whom they referred during the clinical trial<br>5. Require patients in studies to continue to see their primary GI at least once per year for health maintenance (i.e. vaccinations, colonic dysplasia surveillance) |
| Fear of flaring because investigational therapy is ineffective or at too low a dose | 1. Provide data to support rationale for therapy<br>2. Provide data on efficacy in animal models and previous phases<br>3. Provide evidence that dosing will adequately treat target molecule or pathway<br>4. Provide efficacy data in IBD patients (if available) from previous phases of development |
| Fear of adverse events due to investigational therapy | 1. Provide data on safety in humans in previous phases compared to placebo<br>2. Provide data on safety of comparable therapies in the same pathway |
| Concerns about time commitment | 1. Payment to patient for time required, including visits, travel, and data entry<br>2. Provide support for family care – child care, and/or elder care<br>3. Provide support for family absences, including layperson letters to inform friends |
|  | and family of the number of visits, time commitment, and duration<br>4. Provide support for work absences, including letter to inform employer of the number of visits, time commitment, and duration |

Fear related to their illness or treatment appeared to be a major barrier to participation in clinical trials. Some participants expressed that clinical trials conjured up thoughts of a disease that was out of control and needing a “last resort” option. Additionally, there was fear of experiencing a severe flare while in the placebo arm, and a fear that the study medication would not be efficacious or would have significant side effects. Reported opportunities to overcome these fears included better communication to explain the benefits of participating in a clinical trial, and the potential benefits of each trial.

Current disease severity was often mentioned as an important determinant in whether or not a patient would be interested in clinical trials. Many patients said that when their current health situation was bad, they would be more willing to participate in clinical trials. At the same time, being sicker made it more difficult to put in the effort and time needed to participate (e.g., time and travel requirements) and the risks of worsening (e.g., getting a placebo, flaring, adverse events) added concerns that severe flares, hospitalization, and surgery could result from inadequate experimental therapy or placebo treatment. Communication on the length of time a trial medication could take to induce a response is important to set expectations for participants.

The potential adverse effects of drugs or trial participation repeatedly came up as a concern for participants. In short, participants wanted to avoid any adverse effects, particularly if their disease was already active. This finding was supported in a broader study, where the possibility of side effects (mentioned by 43% of adults) was the most frequently mentioned risk associated with clinical trial participation^3^. The likelihood of being in a placebo group was another prominent concern, because participants feared the possibility of a severe flare and the resulting consequences. However, if the trial only had a 20% chance of receiving a placebo participants felt that was tolerable.

The time and travel necessary to participate in a clinical trial also came up multiple times throughout focus group discussions. Patients did not want to spend a great deal of time participating in a trial. This included time spent in individual appointments (specifically lab work), travelling to appointments, frequent treatment appointments, and the duration of the entire trial. In particular, patients did not want to have to spend a lot of time traveling to receive treatment, although greater travel would be considered reasonable for more complex routes of administration (e.g., infusions). The key reasons for not wanting to invest large amounts of time in studies were related to work and family obligations. To overcome this barrier, participants recommended providing comprehensive support to cover their expenses, including reimbursement for their travel and supportive services for family care and flexible appointment scheduling. Patients also expected a high level of professionalism, customer service, and compassion at the facility where they would participate in a trial.

A major topic, discussed multiple times, was the relationship and support with their family and with their doctor, and its influence on participation in clinical trials. The support of family was important, and concerns of family members could be a barrier to participating. Other studies suggest that family support may be an especially important factor in minority populations^2^. Relationships with doctors were more complex. Most patients conveyed a desire for their current or primary GI doctor to support and be aware of their participation in a clinical trial. Many also expected to maintain some degree of communication with this provider about their health status as it related to the trial during and after the trial. Most also wanted their primary GI to be notified of their participation and progress in a trial.

This study is based on a small sample size in five focus groups, with over one quarter having prior trial experience. Despite the small sample, there were several recurring themes across focus groups. The repeated mention of these topics (i.e. fears, disease severity at onset of trial, potential adverse effects of drugs or trial participation, time constraints, and the influence of provider and support network) suggests that they are valid concerns of IBD patients regarding enrolling in clinical trials. It is also possible that with this small sample size, uncommon issues important to IBD patients were not identified or not represented in our sample.

In summary, fear and misunderstanding of clinical trials, engagement with primary gastroenterologists, percentage receiving placebo, time demands, and impact on work and family are barriers to clinical trial participation. Creative solutions to these problems could lead to greater participation in IBD clinical trials and more rapid advancement of new therapies to clinical approval and use. Furthermore, small numbers of participants felt strongly that clinical trials are only for people looking for a last resort to treat their disease, that the ability of potential participants to enter clinical trials was greatly limited by lack of child care, and that traditional media advertising about clinical trials is untrustworthy. These topics need further study to determine how prevalent these attitudes are in the IBD patient population.

## Data Availability

Data sharing not applicable to this article as no datasets were generated or analysed during the current study.

## Acknowledgements

The Center for Information and Study on Clinical Research Participation conducted two focus groups and the Survey Research Institute at Cornell University conducted three focus groups. The Crohn’s & Colitis Foundation supported this study with funding from Biogen MA Inc. and Celgene Corporation.

## Appendix

### Facilitation Guide 2 (used for FG 3-5; FG 1 & 2 was similar)

#### A. General Awareness/Communication of Clinical Trials (30 Min)6:09-6:39

I’d like to start our discussion by asking you about your current knowledge of clinical trials.

1. What do you think of when you hear “clinical trial?”
2. Have you ever become aware of a clinical trial that might be appropriate for you to participate in?
  a. [IF YES] Why did you or did you not follow up on it?
  b. [IF YES] How did you become aware of it?
  c. [IF YES] What did you think of that method as a source of information?
3. Show of hands, how many of you has had your provider talk to you about the opportunity of participating in trials? *Announce number of hands raised*
4. How many of you have participated in a clinical trial before? Announce number of hands raised.
  a. [For those who said NO]: Before you came in today, were you familiar with what a clinical trial is?
  b. [For those who said YES]: *Ask one question at a time and go around in a circle*.
    i. What was the trial for and what motivated you to join the trial?
    ii. How did you hear about the trial (e.g. from your doctor, personal web search, social media, etc.)?
    iii. What was good? What was bad?
    iv. Would you do another one?
    v. Would you share (about available clinical trials) with your friends, family or disease network?
5. What are your impressions of clinical trials for Crohn’s disease/UC in general?
  a. What, if anything, <u>attracts</u> you about participating in a clinical trial for Crohn’s disease/UC?
  b. What, if anything, <u>concerns</u> you about participating in a clinical trial for Crohn’s disease/UC?
6. Do you actively look for information on clinical trials for Crohn’s disease/UC?
  a. Where specifically do you or would you go to find information about potential clinical trials for Crohn’s disease/UC?
    i. Internet search?
    ii. Blogs?
    iii. Social media (Twitter/Facebook/etc.)?
    iv. Doctor? Primary care or specialist?
    v. Patient groups?
    vi. Traditional media? TV/print/radio?
    vii. How easy to access are these sources? Is the information available generally easy to understand or not?
  b. Out of the methods we just talked about, which do you think is the best way to find out about clinical trials?
  c. Who would you want delivering it to you? In other words, who would you trust in getting the information to you? (e.g. Provider, Crohn’s & Colitis Foundation, study staff, other)?

#### B. More information on the trial (25 Min)6:39-7:04

*Okay, now we are going to move on and ask you about what information you would need in order to make an informed decision on whether or not to participate in a clinical trial*.

1. What information would be helpful for you to know about a clinical trial for Crohn’s disease/UC in order to make a decision on whether to participate?
  a. How much time do you think you need to make this decision (if any) after learning about the opportunity?
2. What factors should a recruiter consider when trying to reach you (as a patient) for a potential clinical trial? *(Open-ended, go in a circle again, below are probing questions)*
  a. What is the best way to reach out to you?
  b. What type of communication would you prefer?
3. On a scale of 1-10, where 10 is most important, how important is it that your primary gastroenterologist refers you/supports your involvement?
4. Do you know of any new and recent upcoming clinical trials for Crohn’s disease/UC?
  a. Did you look into them? If not, why not?
5. What message or information about a clinical trial for Crohn’s disease/UC would be most likely to catch your attention?
  a. What would be the best way to deliver this information (ex. email, social media, etc.)?
  b. Is there a specific format that you would like to receive information about a clinical trial for Crohn’s disease/UC (e.g. Visual aid, the internet, radio, phone calls, flyers, etc.)?
  c. Would hearing from another IBD patient who has participated in a clinical trial influence your decision to participate?

#### C. Factors to deciding on trial participation (30 Min) 7:04-7:34

*Now we’d like to ask what factors are important to you when deciding to participate in a clinical trial. Please review the sample clinical trial process that we have put at the table if you haven’t already. I’ll give you a couple of minutes to look over the packet*.

1. Do you know the difference between Phase 2 and Phase 3 trials*?* Would you be more willing to participate in one phase over the other? *Explain if need be:* Phase 1 – tests safety in healthy patients. Phase 2 – safety in patients with IBD, and first look at whether there is a benefit (often testing multiple doses to pick the best dose for phase 3) in a small number of patients. Phase 3 – after efficacy established in phase 2, optimized dose vs. placebo to prove effective in large number of patients
  a. Prior to today, how many of you knew the difference between induction, maintenance, and open label extension phases of an IBD trial? Show of hands. *ANNOUNCE # of hands
2. What are the most important factors in determining if a trial is appropriate? *(Open-ended, don’t read below as it is a probing map, use them if not touched on during discussion)* [Potential factors that can be raised if not done organically]
  a. Drug
    a. Safety profile (potential side effects and risks of treatment found in earlier research)
    b. Efficacy profile (how effective treatment has been found to be in earlier research)
    c. Open label Extension trial (e.g. up to 7 years free drug if it works for you)
    d. Route of administration (pills vs. skin injection vs. IV)
  b. Study process
    a. Phase of study
    b. Treatment arms (e.g. how many people will get placebo)
  c. Physician
    a. Regular physician contact/care
    b. Coordination with your primary GI or PCP
  d. Study Visit Experience
    a. Complexity of trial assessments
    b. Invasiveness of trial assessments – number of colonoscopies, flex sigs, CT scans, MRI scans, stool samples, handling stool, number of biopsies. How close together colonoscopies are
  e. Logistics
    a. Location of trial site/distance in miles
    b. Parking
    c. Number of visits
    d. Need to do visits during 9-5 working hours vs. after hours
  f. Other?
3. Medication regimen
  a. Do you have a preferred method of receiving your treatment?
    a. E.g. Via injection under the skin vs. infusion vs. oral tablets?
  b. What frequency of dosing would you feel comfortable with?
    a. Once a day vs. twice a day vs. every 2 vs. 4 weeks vs. 8 weeks?
  c. Would you participate in a trial if you knew there was a possibility that you could get a placebo? Under what circumstances would you participate in a trial where you could potentially get a placebo?
    a. *Definition if needed: A placebo is an inactive treatment or “sugar pill”. While a placebo may look like the real medical treatment being studied, it does not contain the active medication. Placebos are used to help scientists more clearly understand whether a new treatment is safer and more effective than no treatment at all. You would be randomly (like tossing a dice) assigned to 1 of 2 treatment groups: active or placebo. So for every 3 patients in the trial, 2 will receive the active drug and 1 would receive placebo. The placebo will have no effect on your body and will not harm you nor help you. Neither you, the study doctor, nor the study staff will know whether you are receiving active treatment or placebo. However, in an emergency, your study doctor can find out what treatment you are receiving*.
    b. Does it matter if you can continue on your current medication and add the study medication (this is standard)?
  d. What about induction drug vs. placebo, then opportunity to go to open label if you are not doing well? How long would you be willing to hang in there if not doing well, in order to get active drug later (4w, 6w, 8 w, 10w 12w, 16 w)?
  e. Does the percentage of placebo matter (20%? 16% 33% 50%)? *For facilitator: When you enroll in an induction study, there may bet 2-6 arms, one of which is placebo. Depending on how many non-placebo arms, the percentage of patients getting placebo will vary*.
4. Physician contact/care
  b. Do you have any concern around the relationship with your current provider while you are in a clinical trial (e.g. if they would not be administering your care, etc.)?
  c. Does it matter if your doctor refers you to a clinical trial? Does it matter if your doctor gets notes from each clinical trial visit, so that your doctor knows what is going on?
  d. What if your doctor was not in favor of you participating in a trial? Would that affect your decision to participate? If so, how much influence does your doctor’s approval have on your decision?
5. Location
  a. Does the location of the medical center affect your decision to participate?
  b. How far would you be willing to travel to participate in a clinical trial? [PROBE: What would help alleviate concerns about distance]
  c. Does reimbursement for travel (55 cents per mile), parking, and hotel if farther than 3 hours away, make a difference? Are any other costs a factor for you?
  d. Would you rather participate in a trial in a clinic that does few IBD trials, or in an IBD center that has a lot of experience with clinical trials? How much does this matter?
6. Evaluations
  a. Would you be comfortable participating in a clinical trial for:
    ▪ 3 months
    ▪ 6 months
    ▪ 1 year
    ▪ 2 years
  b. What is your expectation for the length of each study visit?
  c. How long a gap between study visits is expected? E.g. 1 week, 1 month, 2 months, etc.
7. Personal life
  a. Does/would work or personal life get in the way of you being able to participate?
  b. Are/would your loved ones supportive of the idea behind clinical trials?

#### D. What influences your decision to sign-up for a specific trial (20 Min) 7:34-7:54

*Now we’d like to ask about factors that could influence your decision to participate in a clinical trial*.

1. What are the most important factors in signing up for a given trial? [Probe for each – why/how would this effect you?]
  a. Possible side effects?
  b. Time off of school/work?
  c. Complexity of assessments?
  d. Ability to “leave” trial?
  e. Additional treatment available if responding?
  f. Other factors??
2. What are the biggest hang ups of participating in a clinical trial?
  a. Complexity of trial assessments? [Prompts if needed]
    - Invasive procedures?
    - Daily diaries?
    - Blood draws?
    - Other?
  b. Visit frequency?
  c. Time at trial site?
  d. Placebo arm?
3. Is there anything you expect to experience/receive if you participate in a clinical trial for Crohn’s disease/UC?
  a. What is your expectation for number of visits, how frequently the visits would occur?
  b. What is your expectation for how you will be treated at the facility that you choose to participate at?
  c. What is your expectation for post-trial (e.g., continuation of the study medication if it works)? What would make participation in clinical trials easier?
    ▪ [PROBE: extended night or weekend hours/child care/travel assistance to/from appointments/reimbursement of costs for missing work]
4. In general, is there anything that might keep you from participating in clinical trials? [PROBE: Family? Work? Other?]
5. Who from your support environment would you want to help make a decision like this?
  ▪ Doctor?
  ▪ Nurse?
  ▪ Friends/Family?
  ▪ Patient groups?

#### E. Closing comments/suggestions (6 min) 7:54-8:00

1. What are some additional issues or things we need to consider or have not yet discussed?

